# Prosthesis hand grasp control following targeted muscle reinnervation in individuals with transradial amputation

**DOI:** 10.1101/2022.06.03.22275703

**Authors:** Ann M. Simon, Kristi L. Turner, Laura A. Miller, Gregory A. Dumanian, Benjamin K. Potter, Mark D. Beachler, Levi J. Hargrove, Todd A. Kuiken

## Abstract

Transradial amputation is the most common level of major upper limb amputation. Despite the growing availability of multifunctional prosthetic hands, users’ control of these hands and overall functional abilities remain limited. The combination of pattern recognition control and targeted muscle reinnervation (TMR) surgery, an innovative technique where amputated nerves are transferred to reinnervate new muscle targets in the residual limb, has been used to improve prosthesis control of individuals with more proximal upper limb amputation levels (i.e., shoulder disarticulation and transhumeral amputation). The goal of this study was to determine if similar prosthesis control improvements could be seen for individuals with amputations at the transradial level. Participants controlled 3-5 grips with a multi-articulating hand prosthesis under myoelectric pattern recognition control for at least 8 weeks at home pre- and post-TMR surgery. Users gained some significant functional control benefits using a multi-articulating hand prosthesis with pattern recognition at 9-12 months post-TMR surgery. Additionally, a majority of subjects noted an improvement in their residual limb and phantom limb sensations post-TMR. An additional offline EMG analysis showed a decrease in grip classification error post-TMR surgery compared to pre-TMR surgery.

## Introduction

Transradial amputations account for over 40% of major upper limb amputations (1) and greatly affect an individual’s ability to perform functional tasks in daily life. Due to the amount of dexterity lost in the missing hand, single degree of freedom prosthetic hand components, whether more cosmetically shaped hand and fingers or hook-like terminal devices, are used more as a tool than as a replacement (2). Two main factors that have limited improved function with transradial prostheses include the lack of robust, natural control of more than one degree of freedom and effective hand prostheses that can provide more of the prehensile functions of the intact hand (3).

The newest generation of prosthetic hand components addresses one of these factors. More anthropometric, multi-articulating hands with variable design tradeoffs (2,4,5), present users with a larger set of grasp patterns to hold and manipulate objects of various sizes, shapes, and weights. Control options to switch between grips include smartphone applications, buttons mounted directly to the prosthesis, proximity sensors, IMU gesture-based control, and/or myoelectric control (5–8). Even with all these options, the cognitive burden remains high and affects users’ ability to quickly and reliably choose an appropriate grip for activities of daily living. Myoelectric pattern recognition is one way to reduce the cognitive burden of prosthesis control (9), whereby residual limb muscle contractions can be used to train an intent recognition system (10–12). With pattern recognition of hand control, users can make physiologically appropriate muscle contractions to close their phantom hand in various grasp patterns or individual finger motions to control a prosthesis (13–17).

Targeted Muscle Reinnervation (TMR) surgery is an additional significant advancement in the control of multifunction prostheses (18–20). With TMR surgery, an amputated nerve is transferred to a remaining muscle site in the residual limb and, over the course of several months, the muscle is reinnervated sufficiently that EMG signals can be measured using surface electrodes. Reinnervated muscles, especially for more proximal upper limb amputation levels (i.e., shoulder disarticulations and transhumeral amputations) can provide important additional neural control information for improved myoelectric prosthesis control (11,18,21–23). Because the transferred nerves contain both motor and sensory information, target sensory reinnervation is also possible in the residual limb. To improve sensory outcomes, skin over the targeted muscle is denervated and then sensory nerve fibers in the transferred nerves reinnervate this targeted area of skin. When reinnervated skin is touched the individual feels as if the missing hand is touched (24); reinnervated skin can sense touch, temperature, pain and vibration (18). A potential further benefit of TMR surgery is its potential to treat painful neuromas (25); a recent clinical trial showed TMR surgery improved phantom limb pain and trended toward improved residual limb pain compared to conventional neurectomy (26).

For individuals with a transradial amputation, TMR surgery has the potential to restore data from intrinsic hand muscles. The inclusion of neural information from intrinsic hand muscles has been shown to improve hand grasp and finger movement classification (27). For individuals with a transradial amputation to benefit from this additional information via the TMR procedure it would require denervation of extrinsic hand muscle motor points, potentially removing intrinsic muscle data. It is currently unknown to what extent individuals with more distal amputation levels can benefit from improved prosthesis control following TMR surgery in the residual forearm.

All intrinsic hand muscles are innervated by the median and ulnar nerves, thus only two nerve transfers are needed to provide more complete neural information for pattern recognition control. Potential target muscle sites in the residual forearm include flexor digitorum superficialis (FDS), flexor digitorum profundus (FDP), or brachioradialis for the median nerve and flexor carpi ulnaris (FCU) or flexor pollicis longus (FPL) for the ulnar nerve (28–30). These target muscles are superficial and have close proximity to the donor motor nerve. Furthermore, there are other muscles in the forearm that perform similar functions to these muscles so no important function or electromyography (EMG) information should be lost as a result of the transfers.

The goal of this study was to provide an in-depth evaluation of hand grasp control for individuals with a transradial amputation following TMR surgery. We sought to 1) determine if TMR improved functional outcome measures for individuals with transradial amputations, 2) assess improvement in EMG signals during offline analysis, and 3) evaluate user’s subjective representation of their phantom limbs after TMR.

## Methods

Individuals were recruited from across the US and all fittings and study testing were conducted at the Shirley Ryan AbilityLab in Chicago, IL and Walter Reed National Military Medical Center in Bethesda, MD.

Inclusion criteria were: age 18-95, unilateral amputation below the elbow, and demonstrated the ability to use a myoelectric prosthesis. Exclusion criteria were: inability to use a prosthesis, cognitive impairments that would interfere with their understanding of study requirements, or any significant co-morbidity that would preclude completion of the study. The study was approved by both Northwestern University’s and Walter Reed National Military Medical Center’s Institutional Review Boards. All participants provided written informed consent.

Subjects were fitted by a certified and licensed prosthetist with a custom-made myoelectric prosthesis consisting of a multi-articulating hand, the i-limb Ultra Revolution (OSSUR (31)), a passive wrist rotation component, and a COMPLETE CONTROL Gen 1 system (Coapt, LLC (32)). Eight pairs of stainless steel dome electrodes were embedded into a flexible inner liner. Two electrodes pairs were placed over the intended sites for the TMR surgery identified with the help of the surgeon and six electrode pairs were placed over additional residual limb forearm muscles. The intention was to take additional care in placing the electrodes in this desired manner, ensuring data from the TMR regions where included in both pre and post op TMR surgery prosthetic sockets.

The clinically available pattern recognition system (21,33), customized to record usage, allowed users to recalibrate their control at any time by making natural muscle contractions that followed along with a series of pre-programmed grips (34,35). To switch between grips users a sustained a ‘hand open’ muscle contraction signal returning the hand to a natural baseline hand position before performing the natural muscle contraction for the new desired grip. This scheme balanced the potential desire to remain in a grip even if the hand was fully open and only switch to a new grip following a sustained hold open.

### Pre-TMR Home Trial

Individuals participated in therapy with a certified and licensed occupational therapist to learn how to control the multi-articulating hand with pattern recognition. Clinical judgment and user feedback were used to determine the maximum number of grips the user could access. This determination included a discussion of the user’s activities of daily living, demonstration of the 8 available i-limb grips (a subset of all grips clinically available on the i-limb hand (31)), and practice with the hand with pattern recognition control. The selection of 8 grips included lateral, power, thumb precision pinch opened and closed, thumb 3 jaw chuck opened and closed, standard 3 jaw chuck closed, and index point. No alternative methods of changing grips (e.g., grip chips, IMU-based gesture control, smartphone application, etc) were allowed.

Individuals received a minimum of four training sessions of 1-3 hours in length prior to taking the prosthesis home for at least 8-weeks (Pre-TMR). While at home, users had the ability to use the auto-calibration procedure whenever they desired. During auto-calibration users are prompted to follow along and perform muscle contractions in sync with pre-programmed motions of the prosthesis while EMG data is recorded and auto-labeled for each trained grip. Participants were required to check-in regularly with the occupational therapist, maintain an average usage of at least 2 hours per day with the study device, and log the activities they were performing with their prosthesis. Notably, subjects were allowed to use their own prescribed (i.e., non-study) prostheses if they desired, so long as they maintained an average usage of at least 2 hours per day with the study device. This decision was made as their non-study device may have had a powered wrist rotator or different terminal device (e.g., hook) which may have been more suitable for their occupation and/or some of their activities of daily living. Electronic usage data (duration of time the prosthesis was turned on, which grips were used, and the number of times the prosthesis was recalibrated) was recorded on the pattern recognition controller.

At the conclusion of the home trial, participants returned to the lab to download the usage data logged on the controller and to perform a series of outcome measures (36) including: the Southampton Hand Assessment Procedure (SHAP) (37), the Jebsen-Taylor Hand Function Test (38), the Assessment of Capacity for Myoelectric Control (ACMC) (39,40), the Modified Box and Block Test of Manual Dexterity (41), and the Activities Measure for Upper Limb Amputees (AM-ULA) (42). A higher score for the SHAP, ACMC, AM-ULA, and Box and Blocks and a lower score for the Jebsen-Taylor indicate better function and/or control.

### TMR Surgery and Post-TMR Home Trial

Participants then underwent TMR surgery in their residual forearm. The planned surgery involved transferring the remaining ulnar nerve to the flexor carpi ulnaris muscle and the remaining median nerve to either the flexor digitorum superficialis or brachioradialis muscle (28–30). Targeted sensory reinnervation (24) was discussed with each individual prior to surgery and optionally performed for those subjects who consented to this additional procedure.

After surgery, participants were allowed to resume prosthesis use of their own prescribed prosthesis as soon as they were comfortable. After at least six months post-surgery, in order to allow the TMR sites to re-innervate and mature, a prosthetist re-checked socket fit of the study prosthesis. Participants received additional occupational therapy to re-assess their selected grips and practice pattern recognition control. They participated in another 8-week home trial (Post-TMR-1) and in-lab outcome measure testing similar to the pre-TMR home trial. Participants then completed one last home trial, this time for 3 months in duration with no minimum usage requirement. They returned to the lab a final time to perform outcome measures (Post-TMR-2).

From the electronic usage data we calculated average daily wear time as the total number of hours the device was turned on divided by the total number of days the device was turned on. Since the configured grips for each user were selected based on the most functional grips for their own activities of daily living, the time spent in each grip was ranked in order of descending usage and the percentage of time spent in each grip was calculated. To compare outcomes between pre- and post-TMR surgery, we performed a repeated measures analysis of variance (ANOVA) with subject as a random factor, condition (pre-TMR, post-TMR-1, post-TMR-2) as fixed factors. When p-values were found to be less than 0.05, we made multiple comparison with a control condition (pre-TMR) using Dunnett’s method (43). To make comparisons between the home-logged data for wear time, and frequency of recalibration, we used the same model as above, but with only two conditions (pre-TMR and post-TMR-1) as subjects were not required to wear the study device during the post-TMR-2 period of the study.

### 32-channel EMG Data Collection

To further investigate the potential for advanced prosthetic grasp and digit control, a separate EMG data collection was performed following each 8-week home trial pre- and post-TMR surgery. An array of 32 surface bipolar pre-gelled silver/silver chloride EMG electrodes were placed on each subjects’ residual forearm. Individuals were asked to perform various hand muscle contractions including hand open, 9 different grasp patterns (lateral, power, pinch with the remaining fingers opened and closed, 3 jaw chuck with the remaining fingers opened and closed, index point, hook, and tool), 8 different individual finger and thumb movements (thumb flexion/extension, thumb abduction/adduction, index flexion, middle flexion, ring flexion, and pinky flexion), and rest. Muscle contractions were held for 2 seconds and repeated 8 times for a total of 16 seconds per movement.

An offline analysis was performed on these data using a similar pattern recognition system to what was used during home trial. The data were separated into two separate sets of movements: 1) hand open, 9 different grasp patterns, and rest and 2) 8 different individual finger and thumb movements and rest. For each user and each set of movements, a search was performed to find the optimal 4 channels using classification error (i.e., percentage of incorrectly classified movements divided by the total number of classified movements) as the optimization metric; 4 channels represents a more clinically viable system and it is relatively easy to fit this many EMG channels into a socket. Using the 4 optimal channels per user per movement set, a search was performed to evaluate reduction in classification error as additional grips were added to the system. This analysis was performed separately on the pre- and post-TMR data sets.

## Results

Eleven individuals with a unilateral transradial amputation were recruited for this study (Table 1). Seven subjects completed the full study protocol and one subject, TR1 completed all but the final Post-TMR-2 home trial. One of the seven subjects, TR5, had his post-TMR home trial during the spring of 2020 which was affected by the COVID-19 pandemic; his post-TMR home usage was impacted and his outcomes testing was delayed by 5 weeks. During testing he had indicated he had not used the study prosthesis at all during this 5-week delay. Therefore, his data were excluded from the quantitative analyses. Two subjects withdrew prior to having TMR surgery (TR6 was not able to meet the daily usage requirement and TR8 was unreachable during his second home trial and did not return for outcomes testing) and one subject withdrew after TMR surgery (TR9 withdrew prior to the start of the post-TMR 8-week home trial).

**Table 1.** Participant Demographics

| Subject | Gender | Years Post Amputation | Age Range | Etiology | Prescribed Prosthesis |  | Study Participation |  |
| --- | --- | --- | --- | --- | --- | --- | --- | --- |
|  |  |  |  |  | Terminal Device | Myoelectric Control | Home Trial | 32-Chan Analysis |
| TR1 | Male | 0.8 | 41-45 | Left Trauma | Sensor speed hand, Motion Control ETD | Direct Control | Yes+ | Yes |
| TR2 | Male | 1 | 31-35 | Right Trauma | Michelangelo Hand | Direct Control | Yes | Yes |
| TR3 | Male | 1 | 56-60 | Right Trauma | Motion Control ETD | Direct Control | Yes | Yes |
| TR4 | Male | 1.5 | 26-30 | Left Trauma | Bebionic Hand | Coapt Pattern Recognition | Yes | Yes |
| TR5 | Male | 1.5 | 26-30 | Left Trauma | Bebionic Hand | Direct Control | Yes* | Yes* |
| TR6 | Male | 1.5 | 36-40 | Left Trauma | OSSUR i-Limb, | Coapt Pattern Recognition | Withdrawn |  |
| TR7 | Male | 2.9 | 36-40 | Left Trauma | Bebionic Hand, Motion Control ETD | Direct Control | Yes | Yes |
| TR8 | Male | 3.4 | 36-40 | Left Trauma | Bebionic Hand, Motion Control ETD | Direct Control | Withdrawn |  |
| TR9 | Male | 11 | 36-40 | Left Trauma | Bebionic Hand, Ottobock Greifer, Motion Control ETD, Sensor speed hand | Direct Control | Withdrawn |  |
| TR10 | Female | 12 | 46-50 | Right Trauma | Bebionic Hand, Motion Control ETD | Coapt Pattern Recognition | Yes | Yes |
| TR11 | Male | 12.8 | 51-55 | Right Trauma | OSSUR i-Limb, Sensor speed hand | Direct Control | Yes | No |

ETD: electric terminal device
+ TR1 did not complete the Post-TMR-2 home trial
\* TR5's data were excluded from quantitative analysis due to extensive delay in post-TMR testing as a result of the COVID-19 pandemic.

### Prosthesis Control and Usage

All users successfully used the multi-articulating hand prosthesis both before and after TMR surgery. Only minor fitting adjustments were necessary to the socket post-surgery; the location of all the electrodes remained the same for all subjects. Comparing between the pre-TMR and post-TMR 8 week home trials, there were no significant differences between the number of configured grips (p = 0.054), the cumulative hours powered on (p = 0.234), nor the number of recalibration sessions (p = 0.068) (Table 2) (Figure 1). Grip usage was similar between surgery conditions: on average pre-TMR individuals spent 56.1% [6.6] of the time spent in the preferred grip, and 23.4% [3.5] and 16.5% [3.4] of time spent in the 2nd and 3rd most used grips. Post-TMR individuals spent 52.5% [7.2] of the time spent in the preferred grip, and 24.1% [3.6] and 13.6% [2.6] of time spent in the 2nd and 3rd most used grips.

**Table 2.** Pre- vs Post-TMR 8-Week Home Trial Usage Data

| Metric | Pre-TMR | Post-TMR-1 | Post-TMR-2 |
| --- | --- | --- | --- |
| Number of grips configured | 3.7 [0.8] | 4.1 [0.7] | 4.1 [0.7] |
| Home trial length in days | 69.7 [8.9] | 72.6 [10.5] | 119.3 [33.0] |
| Number of days turned on | 51.1 [15.7] | 43.4 [18.2] | 20.8 [11.3] |
| Cumulative hours on | 264.6 [67.1] | 211.4 [57.1] | 67.8 [38.4] |
| Number of calibrations | 33.1 [30.6] | 12.6 [11.1] | 11.2 [7.1] |
| Grip usage as a percent of total time in hand close |  |  |  |
| Most used grip | 54.0 [19.0] | 46.0 [9.5] | 42.4 [11.2] |
| 2<sup>nd</sup> most used grip | 23.8 [10.7] | 27.4 [5.0] | 27.3 [5.1] |
| 3<sup>rd</sup> most used grip | 18.1 [9.2] | 15.5 [5.7] | 18.8 [5.3] |

Values represent mean [standard deviation]

**Figure 1.**
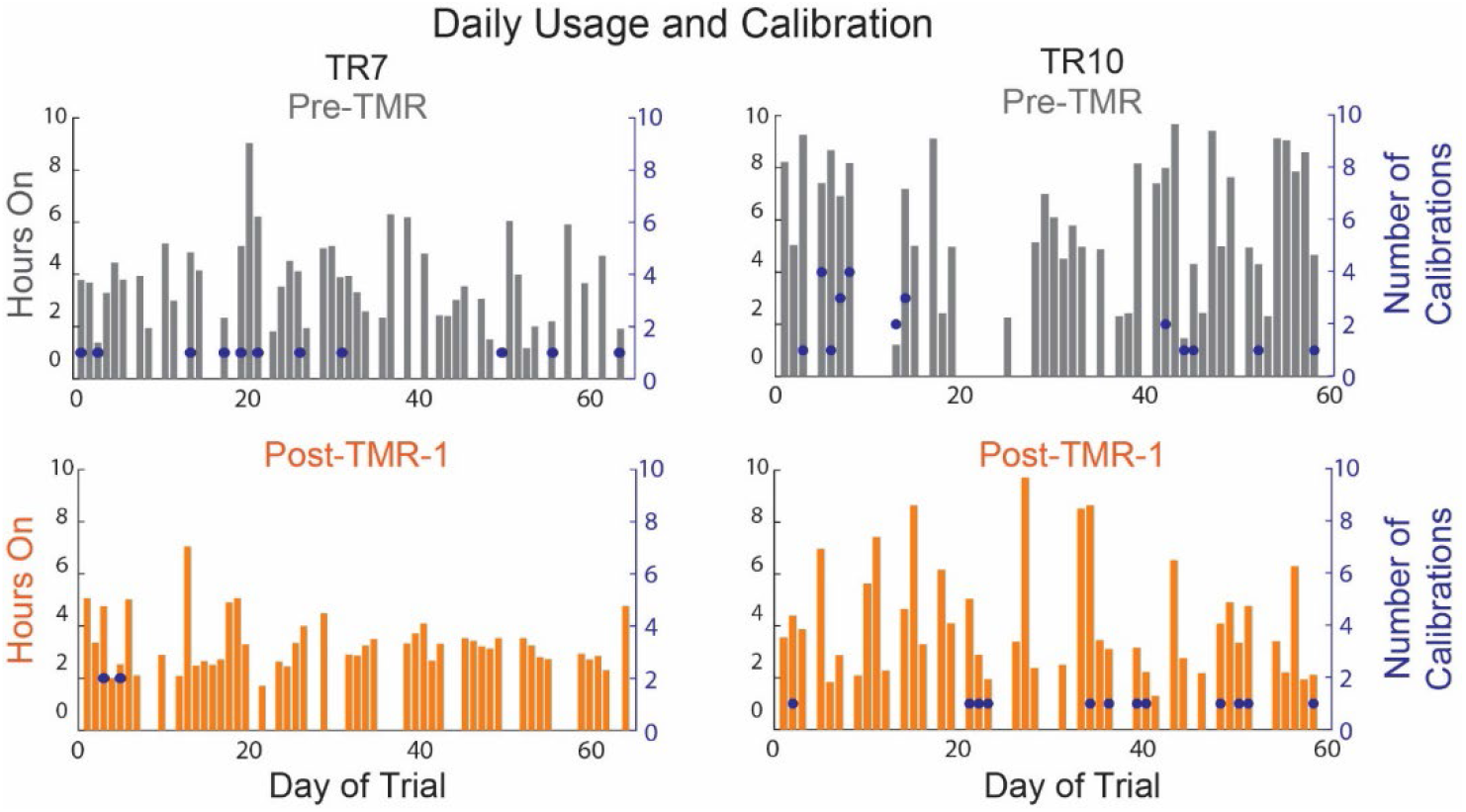
Home usage from two representative participants during the 8-week Pre-TMR (*top*) and 8-week Post-TMR-1 (*bottom*) home trials. Each graph indicates the number of hours the device was turned (bar, with scale on left-hand axis) and the number of calibrations (dot, with scale on right-hand axis) performed for each day of the home trial. Days of no use were logged as 0 hours on the plots.

Users’ index of function score on the SHAP showed a significant average improvement of 12.6 points following the final 3-month post-TMR home trial (Post-TMR-2) compared to pre-TMR (Figure 2, Table 3). Subjects scored significantly lower on the Jebsen-Taylor test of hand function during the Post-TMR-2 condition compared to the Pre-TMR condition. Users transferred significantly more blocks, on average 5.8 more blocks, during the Post-TMR-2 Box and Blocks test compared to Pre-TMR. No significant differences were found between surgical condition for the ACMC or AM-ULA.

**Figure 2.**
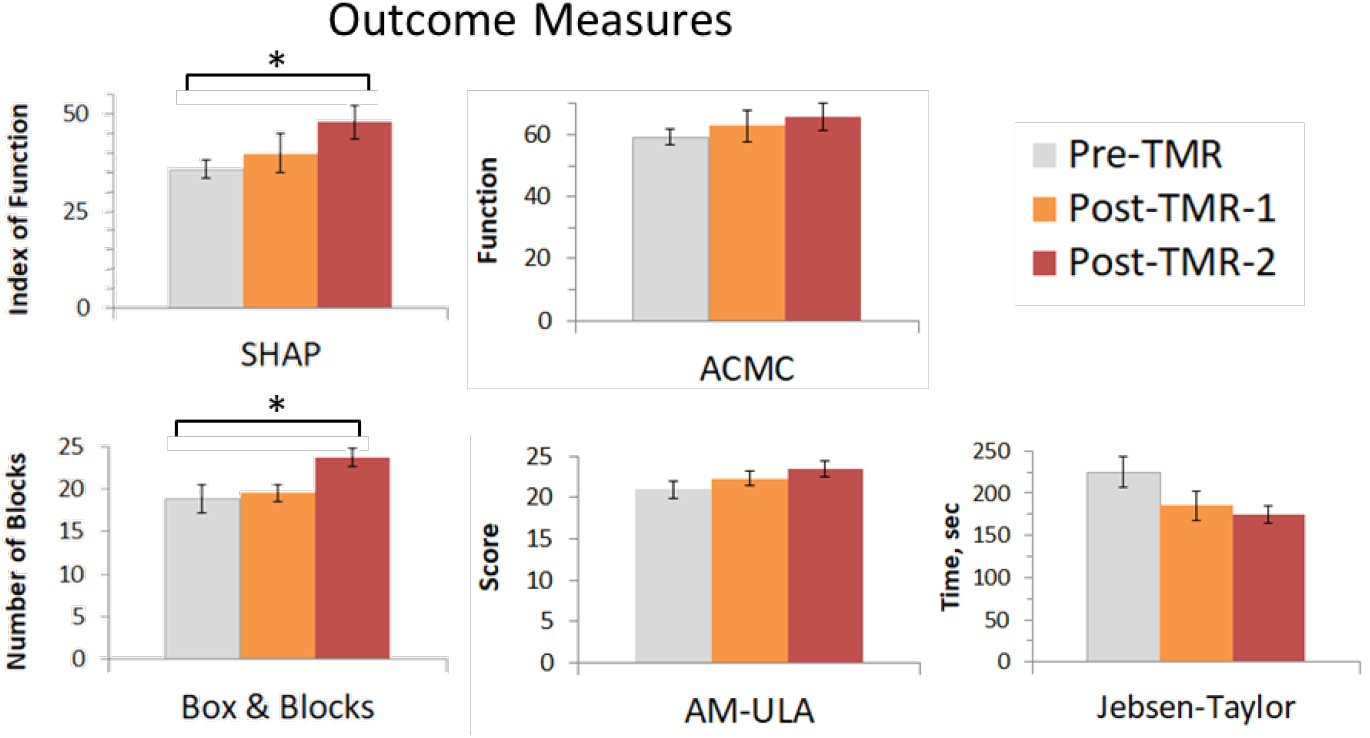
Outcome measures following a Pre-TMR 8-week trial (Pre-TMR, *grey*), a Post-TMR 8-week trial (Post-TMR-1, *orange*), and the final Post-TMR 3 month trial (Post-TMR-2, *red*). The ACMC and Box & Blocks showed significant improvement Post-TMR-2 compared to Pre-TMR (*: p < 0.05).

**Table 3.** Outcome Measures Following Home Trial

| Outcome | Pre-TMR | Post-TMR-1 | Post-TMR-2 | p value |
| --- | --- | --- | --- | --- |
| SHAP, index of function | 35.4 [2.5] | 40.0 [5.1] | 48.0 [4.3] | 0.043 |
| Jebsen-Taylor, seconds | 226.4 [17.7] | 185.1 [16.9] | 174.2 [10.4] | 0.048 |
| ACMC, score | 58.7 [2.1] | 62.8 [2.7] | 62.4 [3.1] | 0.37 |
| AM-ULA, score | 21.1 [1.0] | 22.3 [0.9] | 23.5 [0.9] | 0.14 |
| Box and Blocks, number of blocks | 18.0 [1.6] | 19.5 [1.0] | 23.7 [1.0] | 0.048 |

Values represent mean [standard deviation]

Offline classification errors from the 32-channel EMG data collection using all 32 channels as well as the 4 optimal channels show a slight reduction in error rate Post-TMR compared to Pre-TMR for the grip movement set as well as the individual thumb and finger movement set (Figure 3). When only 4 grips were configured, similar to the amount configured during the home trial portion of this study, classification error rate was 3.2% [0.1] for the 4-channel Pre-TMR data and 2.4% [0.7] for the Post-TMR data. For all 9 grips, error rate was 27.8% [2.7] for the 4-channel Pre-TMR data and 22.7% [3.4] for the 4-channel Post-TMR data.

**Figure 3.**
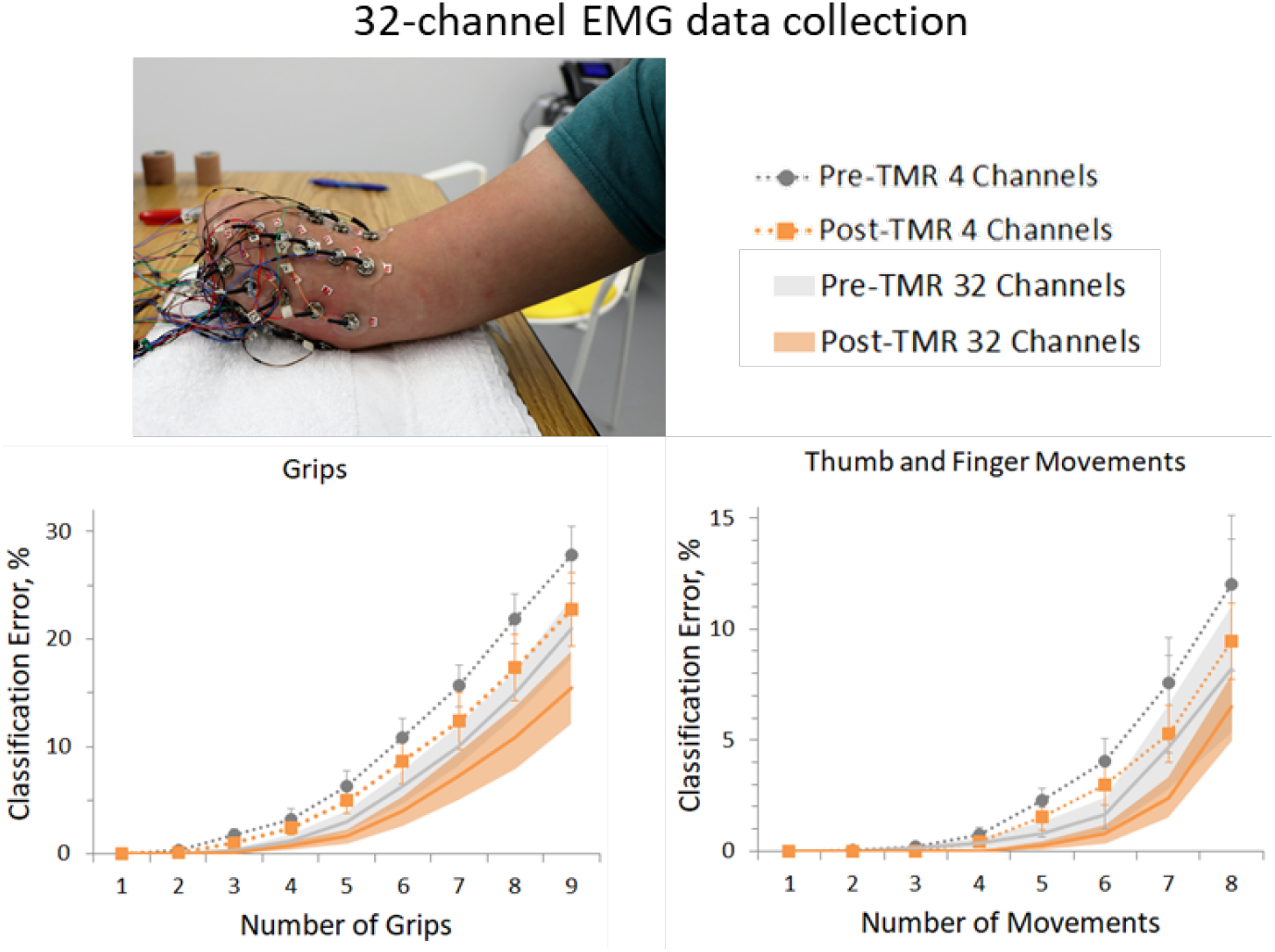
32-channel EMG data collection setup (*top*) and results (*bottom*). Offline classification error rates are plotted for the system with increasing number of grips configured (*left*) and thumb and finger movements (*right*). Colors represent data collected Pre-TMR (*grey*) and Post-TMR (*orange*) optimized for a system using only 4 channels and all 32 channels. Standard deviations for the 4 channels are shown with error bars and for the 32 channel data with shaded bands

### TMR and TSR Qualitative Results

Table 4 summarizes the TMR surgeries for the eight subjects who completed the protocol; two subjects opted to also have TSR. Four subjects noticed an improvement in their phantom limb resting position and/or movement of their phantom hand at the six-month mark or later following TMR surgery. Six subjects (TR2, TR4, TR5, TR7, TR10, TR11) indicated a reduction in overall self-reported pain levels and/or an ability to reduce their overall medications for pain management post-TMR. One subjects (TR3) had no noticeable self-reported change in pain levels and one subject (TR1) who had an increase in residual limb pain post-TMR surgery underwent an additional surgery by a different surgical team after completion of the 8-week post-TMR home trial for neuroma revision of a different nerve. This subject did not complete the Post-TMR-2, 3 month home trial. Four subjects indicated positive improvements post-TMR in the resting position or movement of their phantom limb (Table 4). The two subjects who underwent TSR surgery developed referred sensations from their missing hand in their residual forearm (Figure 4).

**Table 4.** Subject TMR and TSR Results

| Subject | TMR nerve transfers |  | TSR | Phantom Limb Position |  |
| --- | --- | --- | --- | --- | --- |
|  | Median | Ulnar |  | Pre-TMR | Post-TMR |
| TR1 | Brachioradialis | FCU | none | Hand feels “trapped”, minimal ability to move fingers | No noticeable change |
| TR2 | Brachioradialis | FCU | Median sensory nerve to distal aspect of lateral antebrachial cutaneous nerve | Hand feels as if fingers are “wrapped around each other”, cannot move fingers | Fingers rest in a more relaxed position and able to move thumb and fingers slightly |
| TR3 | Brachioradialis | FCU | none | Hand feels mostly “bound up” in a closed fist but sometimes feels “half open” | Resting position remains the same but now can feel minimal thumb and finger movements |
| TR4 | FDS | FCU | none | Hand in constant grip | Able to move thumb and fingers |
| TR5 | FDS | FCU | none | Hand remains in half open fist with minimal ability to move fingers (not thumb) | Resting position remains the same but now pointer and middle finger move together |
| TR7 | Brachioradialis | FCU | none | Able to move thumb and fingers | No change |
| TR10 | Brachioradialis | FCU | Median sensory nerve to end of lateral antebrachial cutaneous nerve | Hand remains in half closed fist, ability to wiggle fingertips | Hand position slightly more open and able to move thumb and fingers more |
| TR11 | FDS | FDP | none | Hand remains in semi-closed fist, very difficult to open fingers | No noticeable change |

FDS: digitorum superficialis; FCU: flexor carpi ulnaris ; FCR: Flexor carpi radialis; FDP: flexor digitorum profundus

**Figure 4.**
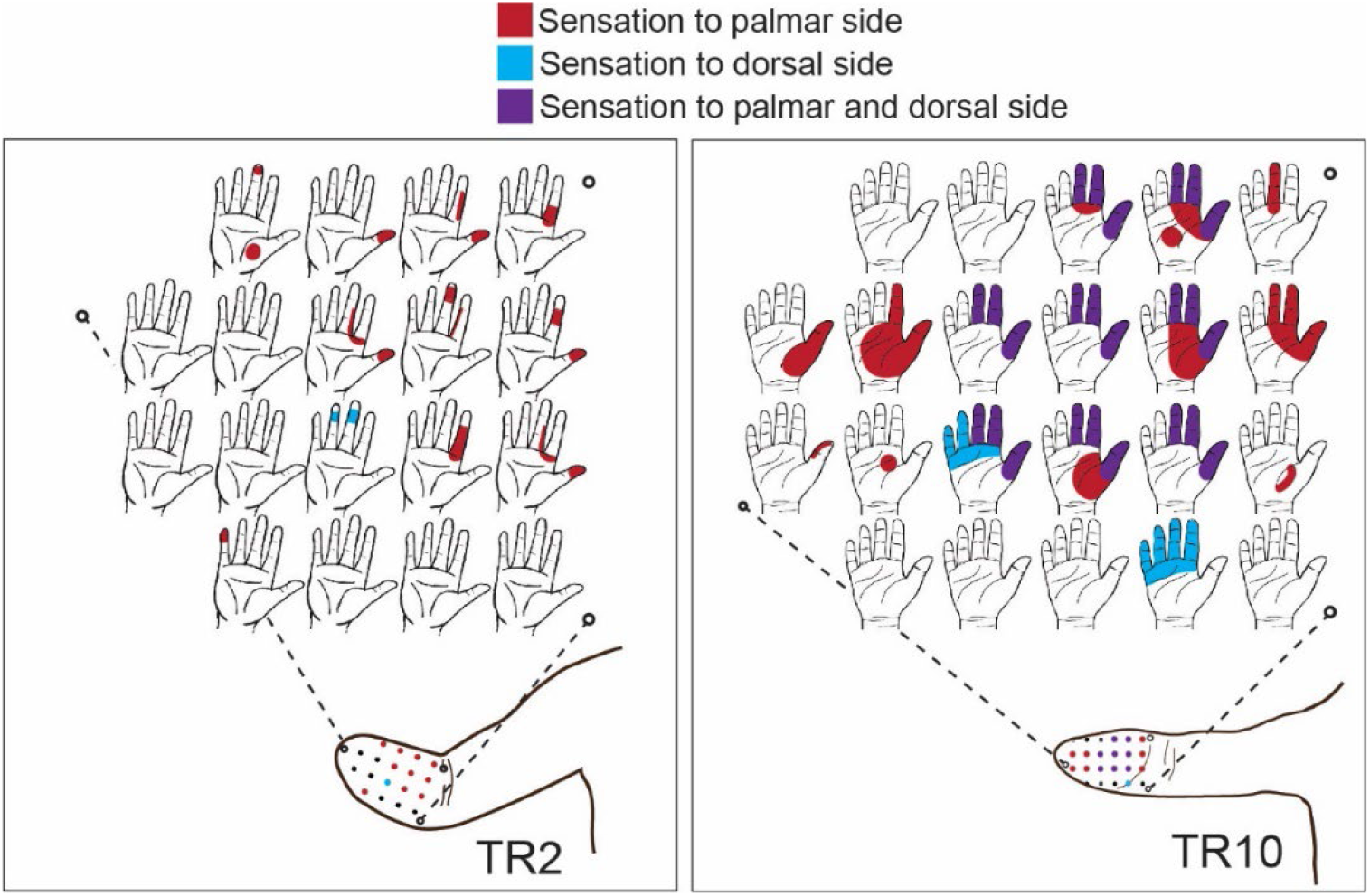
Reinnervated forearm skin of the two subjects who underwent targeted sensory reinnervation. For both individuals, sensations referred from their missing hand were felt in their residual forearm. Referred sensations were localized to either the palmar side (red), the dorsal side (blue), or both (purple).

## Discussion

Our results demonstrate some benefits of TMR surgery for individuals with a transradial amputation. Outcomes data indicate that significant control improvements were seen following the Post-TMR-2 home trial compared to the Pre-TMR home trial for the SHAP, Jebsen-Taylor and Box and Blocks. The SHAP, a timed outcome measure that evaluates unilateral hand functions required for activities of daily living [30], improved by 12.6 points. This increase may indicate that users had an improved ability to function with the prosthesis post-TMR; it is also possible that it is from users gaining a better understanding of the prosthesis or from learning the assessment (45). Users decreased their timed score on the Jebsen-Taylor by an average of 52.2 seconds during the Post-TMR-2 outcomes testing compared to Pre-TMR. The Box and Blocks improvement of 5.8 more blocks is below the minimal clinically important difference at the 90% confidence level of 6.5 blocks (46). With the limited numbers available, potential differences in the ACMC and AM-ULA were not statistically significant between assessment points.

The Post-TMR-2 home trial occurred approximately 9-12 months post-TMR surgery. While it is possible that the targeted muscles took longer than expected to reinnervated, there are other potential explanations. Control improvements may have been due to users having more experience with the multi-articulating hand prosthesis. Due to the nature of the study protocol of users serving as their own baseline for TMR surgery, we were unable to randomize the condition order: 8-week Pre-TMR home trial, 8-week Post-TMR-1 home trial, and 3 month Post-TMR-2 home trial. While additional time with the prosthesis may allow users to better predict hand speed and how the hand moves within each grasp pattern it is important to note that the first two home trials constituted an average of 51 and 48 usage days so the majority of this learning should have already occurred. It is also possible that some of the qualitative TMR surgery outcomes aided users’ consistency and control post-TMR surgery. Reductions in pain, changes in phantom limb sensations, and improved phantom limb awareness may have led to increased repeatability of the muscle contractions necessary for pattern recognition prosthetic control. Further analysis of the calibration data may shed light on whether changes in phantom limb representations and/or long-term pattern recognition use can lead to increased repeatability or separability of EMG signals (47).

The 32-channel EMG analysis also showed a decrease in classification error post-TMR surgery compared to pre-TMR surgery, although these differences were more prominent with a more complex system (i.e., when all 9 grips or all 8 thumb and finger movements were included). Additionally, this analysis showed that four optimally placed EMG channels could provide a highly accurate pattern recognition system: under 10% error for 5 grips pre-TMR or 6 grips Post-TMR and under 20% error for 7 grips pre-TMR or 8 grips post-TMR (Figure 3). However, for this analysis grip muscle patterns were chosen based on which ones were most separable from others in the set as opposed to choosing grips based on what may be most functionally relevant to an individual’s activities of daily living. Therefore, in clinical practice the likelihood is that these systems (or systems that use up to 8 channels) will create slightly higher error rates in grip selection.

The current study saw a smaller improvement in functional prosthesis control than previous studies investigating TMR surgery following more proximal upper limb amputation. This is likely because TMR surgery subsequent to more proximal level amputation allows for access of hand and wrist information where there was none prior. Conversely, following transradial amputation there already exists hand and wrist neural information due to native innervation of the forearm and extrinsic hand muscles. Activation of the newly reinnervated muscles either may not have added considerable neural content or these TMR signals were small and overshadowed by surrounding forearm activation for similar hand movements. It is important to note that denervation of the target muscles used in this study (flexor carpi ulnaris, flexor digitorum superficialis, and brachioradialis muscles) did not lead to worse prosthetic control nor negatively affect residual limb control.

Four subjects described improvements to either the resting position of their phantom hand or the amount of movement they are able to visualize with their phantom. For example, one subject described the resting position of his phantom limb as fingers tightly clenched over one another with a feeling that his fingernails were going through his palm. It had remained that way since his amputation. Post-TMR his phantom hand relaxed such that his hand was more open and in a more relaxed hand position. Additionally, six subjects saw reductions in pain and/or were able to reduce their pain medication levels post-TMR surgery. The one subject who reported no change in pain, however, was no longer taking pain medication that he had been pre-TMR. A recent TMR clinical trial on treatment of neuromas (26) found significant decreases in phantom limb pain and a trend towards reduction in residual limb pain post-TMR surgery compared to conventional treatment of neuromas. One subject who did not experience a reduction in pain, was found to have additional symptomatic neuromas post-TMR surgery due to additional amputated nerve endings that were not targets of the TMR surgery. Evidence shows that post-TMR, the newly cut motor nerves do not typically become symptomatic neuromas (25,26,44).

The current study had some limitations. Because the specific goal was to investigate potential improvements in control due to TMR surgery and the transferred nerves provided additional hand control information, powered wrist control was not included. It is unclear if the addition of a powered wrist would have impacted control or functional outcomes. All participants continued to wear their own, clinically prescribed prosthesis during the home trials; this was allowed as long as they were able to maintain the minimum average usage of 2 hours per day with the study prosthesis. We cannot say definitively what the total prosthesis wear-time (study and non-study devices) was for users. Subjects chose to use their clinically prescribed devices for a variety of reasons including the addition of a powered wrist rotator or the preference to use a different terminal device. As previously mentioned, the order of conditions could not be randomized; therefore, there is the potential for additional learning to have occurred during the post-TMR conditions. Finally, care should be taken while interpreting our statistical results as our lower-than-expected sample size likely impacted the statistical power of the comparisons made. The number of subjects who enrolled and completed the study was lower than planned for due to unexpected subject dropout and the impact of the Covid19 pandemic.

## Conclusion

Transradial amputation is the most common arm amputation and although affected individuals can control wrist and hand movements of a powered prosthesis, functional and reliable control still remains a challenge. The goal of this study was to determine if these individuals could benefit from the additional neural information that TMR surgery provides. Our results show that some significant functional control benefits of using a multi-articulating hand prosthesis were observed 9-12 months post-TMR surgery. Additional qualitative TMR and TSR benefits were observed in some, but not all, enrolled subjects. Future work incorporating both the sensory feedback benefits for individuals with transradial amputation as well as the potential for TMR surgery to result in more repeatable muscle contractions, possibly due to the reduction in pain levels and/or changes to phantom limb sensations, has the potential to increase functional use of many of the clinically available dexterous prosthetic hands.

## Data Availability

All data produced in the present study are available upon reasonable request to the authors

## Acknowledgement

The authors thank Jose Ochoa, Rudhram Gajendran, Michael Stephens, Grant Wang, Matt Mongeon for their technical assistance with this project and Carlee Culver for administrative assistance with the subjects enrolled at Walter Reed National Military Medical Center.

This work was supported by the National Institutes of Health NIH R01HD081525 and 5R01HD094861-04.

## Conflict of Interest Statement

Coapt LLC was launched in 2012 and has a technology transfer and license agreement with the Shirley Ryan AbilityLab for the development of certain control technologies. Todd Kuiken and Levi Hargrove in the Center for Bionic Medicine at Shirley Ryan AbilityLab are responsible for the design, conduct and reporting of this research, and also have financial, management and ownership interests in Coapt LLC, which manufactures the device being tested in this research. These interests have been fully disclosed to Shirley Ryan AbilityLab and Northwestern University, and there is a conflict of interest management plan in place relative to this research study.

## Author Contributions

Conceptualization: TK, GD. Investigation: AS, KT, LM, MB. Methodology - TMR surgery: GD, BP, TK. Formal analysis: AS, KT, LM, LH, TK. Writing – original draft preparation: AS. Writing – review and editing: AS, KT, LM, GD, BP, MB, LH, TK.

## Notes

### Clinical Trial

NCT02349035

### Author Declarations

The Institutional Review Boards of both Northwestern University and Walter Reed National Military Medical Center's Institutional Review Board gave ethical approval for this work. All participants provided written informed consent.

